# A Novel Smart City Based Framework on Perspectives for application of Machine Learning in combatting COVID-19

**DOI:** 10.1101/2020.05.18.20105577

**Authors:** Absalom E. Ezugwu, Ibrahim A.T. Hashem, Olaide N. Oyelade, Haruna Chiroma, Mohammed A. Al-Garadi, Idris N. Abdullahi, Olumuyiwa Otegbeye, Amit K. Shukla, Mubarak Almutari

**Affiliations:** School of Computer Science, University of KwaZulu-Natal, Pietermaritzburg, KwaZulu-Natal South Africa; College of Computer Science, University of Hafr Al Batin, Saudi Arabia; Future Technology Research Center, National Yunlin University of Science and Technology, Taiwan; Department of Biomedical Informatics, Emory University, Atlanta, USA; Department of Medical Laboratory Science, College of Medical Sciences, Ahmadu Bello University, Zaria, Nigeria; IRISA Laboratory, ENSSAT, University of Rennes 1, France

**Author notes:** **Corresponding Authors** Absalom E. Ezugwu, School of Mathematics, Statistics, and Computer Science, University of KwaZulu-Natal, Pietermaritzburg, South Africa;. Haruna Chiroma, College of Computer Science, University of Hafr Al Batin, Saudi Arabia;.

**Keywords:** COVID-19, Drugs discovery, Machine Learning Algorithms, Novel Coronavirus, Smart Cities

## Abstract

The spread of COVID-19 across the world continues as efforts are being made from multi-dimension to curtail its spread and provide treatment. The COVID-19 triggered partial and full lockdown across the globe in an effort to prevent its spread. COVID-19 causes serious fatalities with United States of America recording over 3,000 deaths within 24 hours, the highest in the world for a single day and as of October 2020 has recorded a total of 270,642 death toll. In this paper, we present a novel framework which intelligently combines machine learning models and internet of things (IoT) technology specific in combatting COVID-19 in smart cities. The purpose of the study is to promote the interoperability of machine learning algorithms with IoT technology in interacting with a population and its environment with the aim of curtailing COVID-19. Furthermore, the study also investigates and discusses some solution frameworks, which can generate, capture, store and analyze data using machine learning algorithms. These algorithms are able to detect, prevent, and trace the spread of COVID-19, and provide better understanding of the virus in smart cities. Similarly, the study outlined case studies on the application of machine learning to help in the fight against COVID-19 in hospitals across the world. The framework proposed in the study is a comprehensive presentation on the major components needed for an integration of machine learning approach with other AI-based solutions. Finally, the machine learning framework presented in this study has the potential to help national healthcare systems in curtailing the COVID-19 pandemic in smart cities. In addition, the proposed framework is poised as a point for generating research interests which will yield outcomes capable of been integrated to form an improved framework.

## 1. Introduction

For over 4 months, the novel coronavirus that was later named Severe Acute Respiratory Syndrome Coronavirus 2 (SARSCoV-2) has caused unprecedented Coronavirus Disease 2019 (COVID-19) all over the world. The first human to human transmission of COVID-19 was first reported to World Health Organization (WHO) on 30th December, 2019. Thereafter, several retrospective studies revealed that many COVID-19 patients started showing pneumonia symptoms in early December (Zhang, 2020), (Xiaopei Duan, 2020), (Yuanyuan Dong, 2020). Even though, there are scientific controversies and theories over the date and origin of the SARS-CoV-2, it is widely proven that the novel coronavirus originated from the Wuhan, China (Hao, Zhong, Song, Fan, & Li, 2020). Genomic sequences of the early isolates of SARS-CoV-2 from infected patients in Wuhan showed over 88% nucleotide homology with two bat like SARS coronaviruses. Hence, strongly pointing towards zoonotic source and indeed bats serve as reservoir host of the SARS-CoV-2 (Hao et al., 2020). Currently, there are ongoing searches for the possible intermediate host, which might have aided the transmission of the virus to human. The SARS-CoV-2 are droplet borne pathogen which gets in contact with humans when they are exposed to oral or nasal secretions of clinically symptomatically or asymptomatic infected persons (ECDC, 2020).

SARS-CoV-2 has tropism to cells and tissues that express the angiotensin converting enzyme 2. These receptors are mainly found in the respiratory tracts and limited to the extent of the kidney, heart and gastrointestinal tract. The virus dock onto the receptor by means of receptor binding domain (RBD) of its spike glycoprotein. This represent the first step of viral replication and pathogenesis (Rothan & Byrareddy, 2020). As the virus replicates in the respiratory tract, it’s provokes respiratory symptoms which mainly include dry cough, difficulty in breathing and sore throat. Then, it disseminates through the blood to other tissues and organs causing viremia and high fever. Hence, these symptoms together with others such as body weakness and pains represent the cardinal clinical symptoms of COVID-19 (Lin, Lu, Cao, & Li, 2020).

Majority of the SARS-CoV-2 infected persons remain asymptomatic and infection being self-limiting. However, some 5% of infected persons suffer severe COVID-19 (Rothan & Byrareddy, 2020). Major determining factor for severe and fatal COVID-19 include old age (>60 years), underlying cardiovascular, immunological, metabolic and respiratory comorbidities. Based on available scientific reports, the transmission of SARS-CoV-2 revolves around human, animals and the environment (Chakraborty & Maity, 2020). For now, preserving human life and health security is the major concern of most countries and territories. Hence, prompted the legislation, implementation and enforcement of adequate infection prevention and control measures for high priority pathogen (WHO, 2019). Therefore, combatting the COVID-19 need better understanding.

For better understanding of the COVID-19 pattern of spread, accurate and speed of diagnoses, development of new therapeutic methods, identification of the most susceptible people according their distinct genetic and physiological characteristics, machine learning algorithms are required to analyze the COVID-19 large scale datasets (Alimadadi et al., 2020).As a result of that, attempts have been made by scholars to apply machine learning algorithms in combating the COVID-19 in from different perspective. For example, drugs discovery targeting COVID-19 is proposed in (Ge et al., 2020). Machine learning approach in CRISPR based COVID-19 surveillance using genome is proposed in (Metsky, Freije, Kosoko-Thoroddsen, Sabeti, & Myhrvold, 2020).The classification of novel pathogens for the COVID-19 is presented in (Randhawa et al., 2020). Automated deep learning COVID-19 patient detection and monitoring system is reported in (Gozes et al., 2020; Oyelade and Ezugwu, 2020). Machine learning approach is applied to develop system for creating awareness about wash to contain the COVID-19 (Pandey, Gautam, Bhagat, & Sethi, 2020).Generative network is used for the design of COVID-19 3C-like protease inhibitors (Zhavoronkov et al., 2020).The survival of severe COVID-19 patients was predicted based on machine learning approach by (Yan et al., 2020). Deep learning based system for quantifying the volume of lung infections is reported in (Yan et al., 2020).

However, the major issue with the previous studies is that each of the study focuses on a particular aspect of combatting the COVID-19 pandemic leaving other critical aspect that are required to fight the COVID-19 pandemic. (Spectrum, 2020) reported that hospital emergency rooms across the globe are experiencing floods of people infected with COVID-19 for urgent treatment. As a result of that, unprecedented influx of COVID-19 patient into the emergency rooms is experienced, doctors are grappling with the problem of patient’s triage. Therefore, struggling to decide the COVID-19 patients that require intensive care. The condition of the patient lungs has to be assessed by doctors and nurses. However, volunteer doctors and nurses without pulmonary training can’t assess the patient lungs. At the peak of the COVID-19 crises in Italy, doctors were faced with a serious problem of taken decision on the patient that should be given much needed assistance. In view of these difficult COVID-19 cases facing doctors and nurses, a machine learning system can help to provide clinical decision support to play critical role in the COVID-19 crises, assist hospitals to be fully functional and keep COVID-19 patients alive.

A framework with multiple dimension to combat the COVID-19 in different front can provide better automated solutions to the fight against COVID-19. A COVID-19 pandemic involve a lot of measures to combat it via machine learning such as predicting COVID-19 vaccine immunogenicity, detecting COVID-19 severity, predicting COVID-19 mortality, COVID-19 resource allocation, COVID-19 drugs discovery, COVID-19 contact tracing, social distance, detecting wearing of mask, detecting COVID-19 patients requiring ventilator, predicting COVID-19 patient that is beyond medical intervention and triage COVID-19 patients. All these can be integrated into a single framework to work automatically within a city. Therefore, smart city can be repurposed to combat the COVID-19 by applying framework with multiple measures to fight the COVID-19. To provide smart applications in the smart cities, many technologies are connected together including sensor networks, broadband communications service, sensor devices, wireless sensor networks, internet, and cloud services. Sullivan (2008) pointed out that smart cities in 2020 will be embedded with smart structures such as the smart healthcare, smart security, smart mobility, smart building, smart governance, smart citizens, smart infrastructure, smart technology, smart energy and smart education.

To the best of our knowledge, stands out from other similar existing frameworks, since the study presents the most comprehensive integration of components perceived to integrate well with machine learning to fight COVID-19 from multiple dimension automatically in smart cities reported in the literature. The framework can address the unique challenges in fighting COVID-19 to ease the work of healthcare workers in saving life’s and provide guide for real world execution in smart cities. (Yu, Wang, Liu, & Zomaya, 2016) pointed out that exploring theory is critical for providing guide for effective applications.

The following research questions presents gap in literature motivating the need for this study:

i. Considering the high volume and the raw nature of data generated from internet connected sensory devices, how can machine learning models be well adopted and adapted in inferring higher level information?
ii. What are the challenge associated with the interoperability of IoT technologies with machine learning algorithms in collecting and analyzing COVID-19 data?
iii. Could the higher level information be repurposed in supporting applications (such as detection, prevention, contact tracing, alert-level dissemination system) designed in combating COVID-19?
iv. How can a computational solution based on robust framework be applied to evaluate the environmental and social weaknesses of a city in containing new COVID-19 outbreaks?

In this paper, we propose a solution framework integrated with machine learning to combat COVID-19 in Smart Cities from multiple dimensions. The novelty of the study lies not only in the essential elements of the framework, but how its elements like physical structures, institutions, policy makers, medics, ICT, IoT, big data, and machine learning algorithms seamlessly integrates to manage all kinds of applications and devices.

The other sections of the paper are organized as follows: Section 2 presents the background information about fundamental concepts considered in the study, Section 3 presents the proposed solution framework for combatting COVID-19 in smart city from multiple front, Section 4 presents the applications of machine learning in fighting COVID-19 pandemic in smart cities, Section 5 discusses the outcome of the study by outlining case studies on combatting COVID-19 via machine learning in smart city, before the concluding remarks in Section 6.

## 2. Preliminaries: Overview of the Novel Coronavirus Diseases and Smart Cities

This section presents interesting overview coverage on the fundamental concepts, components, and related works for which the study is focused. This general overview allows for the conceptualization of the framework proposed in the subsequent sections. Our review details on the coronavirus disease and its clinical manifestation, and also on the concept of smart cities as it relates with IoT. In addition, a background knowledge of machine learning and its associated algorithms are discussed.

### 2.1 Novel Coronavirus Diseases: background and cardinal clinical features

In this sub-section, attempt is made to present fundamental knowledge about the novel coronavirus disease ranging from background information to clinical features which this study considers relevant for the implementation of the proposed framework

#### 2.1.1 Background of novel coronavirus disease

Out of the seven Coronaviruses, SARS-CoV-2 is the third highly pathogenic coronaviruses that has afflicted human race. As the incidence and fatality rates of SARS-CoV-2 infection, the etiological agent of COVID-19, continues to rise across more than 210 countries and territories, several preventive and control measures have been adopted to halt the spread of the SARS-CoV-2 and minimize COVID-19 associated death. As at 7:30AM GMT+1, 27^th^ April 2020, there were over 3 million global confirmed cases of SARS-CoV-2 infection with case fatality rate (CFR) of over 7.0% (Worldometer, 2020). European and American countries appeared to have the worst CFRs associated with COVID-19 and least in Africa. Although, there is no categorical explanation for this variation. Several observers have attributed the low incidence rate of COVID-19 in Sub-Saharan Africa to under-diagnosis probably due to inadequate molecular diagnostic capacity. But variation in the genetics, strains, viral proteins mutations and host immune response could have contributed to SARS-CoV-2 virulence and pathogenesis (van Dorp et al., 2020).

Although, there have been controversies on the origin of SARS-CoV-2. However, several studies have been able to trace the source of this infection to zoonotic origin where COVID-19 patients were exposed to an animal market in Wuhan where live animals were sold. Subsequently, efforts have been made to search for a reservoir host and/or intermediate hosts of SARS-CoV-2 from which the infection might have spread to humans. Initially, two snake species were identified as the possible reservoir hosts of SARS-CoV-2. However, the only consistently identified SARS-CoV-2 reservoirs are mammals and bats (Bassetti et al., 2020). Particularly, genomic sequencing of some SARS-CoV-2 isolates show 88% nucleotide homology with two bat-derived-SARS-like CoVs (Lu et al., 2020). Thus, indicating bats as the most likely reservoir hosts for SARS-CoV-2 (Lu et al. 2020).

The principal transmission mode of the SARS-Co-V-2 is through droplets emanating from the respiratory system. These droplets are communicated through coughing and sneezing which uninfected persons who when in contact with it them, contract the disease. These therefore inform on the second principal transmission mode which is contact (Wang et al. 2020). Although younger persons may exhibit some high level of immunity to the disease, studies have however shown that people of all ages are at risk of contracting the disease. The aged who have had contact with infected persons or surfaces often quickly progress to acute respiratory distress syndrome (ARDS) and multiple organ failure. Infected younger persons may stagger along with mild syndrome like fever, fatigue and dry cough and only small percentage cases degenerates quickly as seen in the elderly. The level of propagation of the disease in a city or population is often evaluated using fatality rates and reproduction number (popularly referred to as R_0_ value) and recently, the index c value (Coccia 2020). The effect this propagation has spilled over into social-economic problems which are directly as a result of contracting GDP growth of countries (Coccia 2021).

#### 2.1.2 Clinical features of novel coronavirus disease

Virologically, SARS-CoV-2 is a single stranded RNA virus with positive polarity and variable open reading frames (ORFs) (Cui, Li, & Shi, 2019). It has been shown that two-third of SARS-CoV-2 genome are located within the 1st ORF which translates the pp1a and pp1ab polyproteins. These polyproteins encode 16 non-structural proteins (Cui et al., 2019). Conversely, the remaining ORFs code viral structural and accessory proteins of SARS-CoV-2. The remaining one third genome codes the nucleocapsid (N) protein, spike (S) glycoprotein, matrix (M) protein, and small envelope (E) protein of SARS-CoV-2. Out of these 4 proteins, the S glycoprotein is key because of it is role in host cells attachment and pathogenesis of COVID-19. This protein alongside the viral RNA dependent RNA Polymerase (RdRP) have largely been utilized in the synthesis of primers and antigens for molecular and serological tests of SARS-CoV-2 infection, respectively (WHO, 2020).

Indeed, RNA viruses including SAR-CoV-2 have high mutation rates which are significantly correlated with enhanced virulence and evolvability (Duffy, 2018). At proteomic level, amino acid substitutions have been reported in NSP2, NSP3 and S protein (Wu et al., 2020). Interestingly, another study suggested that NSP2 and NSP3 mutations play significant role in virulence and differentiation mechanism of SARS-CoV-2 (Angeletti et al., 2020). Of interest, is the mutation in S-protein. This has made scientists explore the possible differences of the host tropism and transmission rate of SARS-CoV-2. Worthy to note is the NSP2 and NSP3 mutations in SARS-CoV-2 isolated from many COVID-19 patients in China (Angeletti et al., 2020). These have caused scientists to embark on genomic surveillance of SARS-CoV-2 in order to determine the correlation of these mutations to virulence diversity and their implications on reinfection, immunity and vaccines development (Liu et al., 2020).

Based on available genetic analysis, SARS-CoV-2 is related to SARS-CoV-1 and an early mathematical modelling report revealed an R_0_ of 2 - 3 for SARS-CoV-2 (Yan et al., 2020). This could possibly explain why SARS-CoV-2 is more contagious than SARS-CoV-1 and MERS-CoV which are endemic in certain countries. Infection rate can be referred to as the R_0_, a measure of the transmissibility of SARS-CoV-2. R_0_ predicts how many people an infected person could transmit SARS-CoV-2 in a population with no prior immunity to the pathogen. Generally, the higher the R_0_, the more contagious the pathogen. An R_0_ of < 1 means that the outbreak dies out while R_0_> 1 means the infection will continue to spread (Prompetchara, Ketloy, & Palaga, 2020).

First, it is worthy to note that a single SARS-CoV-2 infected individual has the ability to infect approximately three uninfected persons (Kampf, Todt, Pfaender, & Steinmann, 2020). This occurs through the nano-particles of respiratory droplets which can spread, contaminate surfaces and hands where they remain stable for hours (Kampf et al., 2020). The hands now become a mechanical vector. Thus, a potential site to terminate and prevent it from invading the body. However, if the virus is not eliminated at this stage, it can move towards it is predilection site (i.e. cells of the lungs) where it uses it is spikes to dock and attach angiotensin-converting enzyme-2 (ACE-2) as receptors to gain access into epithelial cells of the respiratory tract. At this stage, SARS-CoV-2 compromises innate lung immunity (Kampf et al., 2020). It then takes advantage of these cells as replication site. The virus regenerates and sheds by disassembling itself and utilizing the machinery of the alveoli cells, precisely Golgi apparatus, to reproduce and repackage itself (Kampf et al., 2020).

The SARS-CoV-2 exists in such a way that as it replicates and disrupts the protective function of the ACE-2 receptor which induces the process of fibrosis (scarring). It has been shown that patients with fatalities associated with SARS-CoV-2 present with a characteristic ground glass effect in their lungs and this sequela impedes efficient oxygenation. As the body tries to compensate for this deficiency, it gradually results to Severe Acute Respiratory Syndrome (SARS), where it becomes impossible for the respiratory system to make available oxygen to the rest of the body (hypoxia) (Kampf et al., 2020). This ultimately results in multiple organ failure. Based on available clinical data, those susceptible to contracting severe form of SARSCoV-2 infection include the elderly (>60 years) population, persons with underlying disease conditions (cardiovascular, metabolic, respiratory and immunological disorders) (Lippi and Plebiani, 2020).

When susceptible individuals get infected by SARS-CoV-2, its either the infected person remain asymptomatic (no apparent illness) or symptomatic. If symptomatic, the disease passes through three stages of severity. At the early infection stage (Stage I), patients present with mild clinical symptoms which includes dry cough, diarrhea, fever and headache. This could last for 3-5 days. These are usually accompanied by lymphopenia (low white blood cell counts), elevated prothrombin time, D-dimer and mild increase in Lactose Dehydrogenase (LDH). Majority (98%) of SARS-CoV-2 infected patients remains at this stage and eventually get cured. However, for those with underlying medical disorders, they may proceed to stage II (Pulmonary Phase), which is predominantly characterized by Shortness of breath and hypoxia (inadequate oxygen supply to the body). This could last from 5 days to 3 weeks). At this stage, patients display abnormal chest radiograph, Transaminitis and declined procalcitonin level. Very few patients (2%) proceed to the severe stage of COVID-19. The stage II (hyper-inflammation phase) is largely characterized by acute respiratory distress syndrome (ARDS), severe inflammatory response syndrome (SIRS), shock and cardiac failure. Majority of patients at this stage eventually die. At this stage COVID-19 patients experience significantly high blood inflammatory markers such as elevated C-reactive protein (CRP), Interleukin-6 (IL-6), D-dimer and ferritin. In addition, affected patients present with increased blood level of Cardiac markers especially troponin and N-terminal (NT)-pro hormone B-type natriuretic peptide (NT-proBNP) (Gao et al., 2020).

Diagnostically, the use of viral culture for establishing acute COVID-19 diagnosis is not practicable due to its long turnaround time (3 days) for SARS-CoV-2 to cause obvious cytopathic effects (CPE) on Vero E6 cells. In addition, isolation of SARS-CoV-2 is laborious and requires biosafety level-3 (BSL-3) facilities which are unavailable in most healthcare centers, especially in developing countries. So far, all available serum antigen (such as the S-glycoprotein) and antibody (IgA, IgM and IgG) detection tests have not been validated by the WHO. However, it has been suggested that serological assays could assist in the analysis of an ongoing SARS-CoV-2 outbreak and retrospective evaluation of the incidence rate of an outbreak (WHO, 2019). In some instances, where epidemiological data of suspected cases correlates to SARS-CoV-2 infection, the demonstration of fourfold rising antibody titer between acute and convalescent phase sera could support diagnosis of COVID-19 when RT-PCR results are negative (WHO, 2019). In addition, it has been revealed that a significant proportion of COVID-19 patients had tested RT-PCR negative despite having suitable clinical features and radiologic findings highly suspicious of SARS-CoV-2 infection (Xiao, Wu, & Liu, 2020). In most cases, these are termed false negatives which could have been due to wrong sampling where SARS-CoV-2 might have been present in the lower respiratory tracts rather than upper respiratory samples often collected during laboratory diagnosis. Hence, this poses a challenge in the proper evaluation of SARS-CoV-2 symptomatic patients (Li et al., 2020).

### 2.2 Rudiments of smart cities and machine learning

This section discussed the concept of smart city including case studies and provide brief explanation about machine learning. This can help readers new in the domain to comprehend the concept of smart city and machine learning.

#### 2.2.1 Smart cities

The universally accepted standard definition of a “smart city” is not yet in existence. In other words, there is no existing standard definition of “smart city”. However, a smart city, is a city that encourages the prudent utilization of quality resource management as well as the provision of services within a limited time. The information and communication technology (ICT) is one of the major component and integrate element in the smart city projects. The operations in smart city cannot be achieved with ICT in isolation. The state-of-the-art view of city development resulted to the new concept of smart city model. The quality and scale of cities have grown significantly as a result of urbanization after the industrial revolution. The expansion in the urbanization have prompted many challenges including (Smart & Cooperation, 2014):

i. Large scale consumption of resources
ii. The degradation of the environment
iii. Unfair widening of gap between the rich and the poor

The smart city has the potentials for helping the cities to achieve sustainability such as high level efficiency, high economy, and improved standard of living for people and beautiful city environment. There are many criteria to assess the smartness of a city as proposed by organizations. These criteria include all or some of the following: smart energy production and conservation, smart mobility, smart economy, smart living, ICT economics, smart environment, smart governance, standard of living and smart society (Smart & Cooperation, 2014). To put smart city in its proper position, smart city is the combination of very large ranges of services that is required by a city and the need to offer the services in such a way that it complies with the current administration requirement through the use of state-of-the-art technology.

#### 2.2.2 Case studies of smart cities

To buttress on the potentiality of smart cities in providing critical support to its population, we briefly review three case studies of application of IoT technology to some cities namely Kuala Lumpur, Copenhagen, and Stockholm.

##### A. Case Study 1: Kuala Lumpur, Malaysia

In Kuala Lumpur, there are many projects in relation to smart city. Many of the projects are in use to optimally utilized the resources in Kuala Lumpur. To reduce traffic congestion, many innovations are implemented in the transport sector. For example, flight check in can be done using smart phones, train and buses schedules are monitored through electronic board place in the train and bus stations in Kuala Lumpur. Smart cards referred to as “touch and go” are commonly used in Kuala Lumpur to avoid long queue in purchasing bus or train ticket. An application referred to as “GrabCar” is used for booking a cap and track the position and estimated time of arrival of the cap to the pick-up position. The weather temperature for the day in different locations in Kuala Lumpur can be monitored through the smartphones. None smoking areas are embedded with sensors to trigger alarm in case of smoking cigarate in the none smoking zone. For the environment, the public transportation mainly used gas for fueling their vehicles to avoid pollution of the environment. Many of the electric vehicles in Kuala Lumpur are hybrid (petrol + electric) which enable switching the engine from electric to petrol and back to electric.

##### B. Case Study 2: Copenhagen, Denmark

In Copenhagen, numerous projects of smart city were analyzed from 2 perspectives, namely, success factors and the economics. The Boyd Cohen list of smart cities in Europe indicated that Copenhagen is ranked number 8. Copenhagen have the vision of becoming the world pioneer carbon-neutral capital by the years 2025. As such, the Copenhagen city is presently implementing innovations in the field of transportation, waste, water, heating and sources of alternative energy to support the target vision of the city by 2025 and enhance sustainability. The Copenhagen has currently expanding its cycle network lanes. The cycle lane is embedded in the broad concept so as to enhance traffic in the Copenhagen city (Catriona Manville et al., 2014).

##### C. Case study 3: Stockholm, Sweden

The Stockholm is currently implementing smart management and applications to ease traffic and issues related to environment. In Stockholm, several waste collecting vehicles are deployed in the city for the management of waste. Despite the efforts put in place, the Stockholm city is facing challenges of waste transport and city traffic. Therefore, half a million entries of waste fractions, weights, and locations were comprehended. The large amount of data about the waste were used for the collection and analysis of waste management collection data. Thus, a shared waste management vehicle fleet is suggested as a solution for the waste problem identified (Shahrokni, Van der Heijde, Lazarevic, & Brandt, 2014).

### 2.3 Machine Learning

Machine learning is a field of science that centers on how a computer learns on data (Abu-Mostafa, Magdon-Ismail, & Lin, 2012). According to (Portugal, Alencar, & Cowan, 2018), “Machine is an algorithm that uses a computer to simulate human learning and allows computers to identify and acquire knowledge from the real-world, and improve the performance of some tasks based on this new knowledge”. Machine learning, a sub-discipline in artificial intelligence, cuts across many fields of studies that correlates with data mining, pattern recognition, computer science (theoretical), and statistics (Deo, 2015). In statistics, it seeks to learn the relationship that exists in data whereas, in computer science, it emphasizes the effectiveness of the computational algorithm. Machine learning research in computer science examines the algorithm that it can learn from and produces a prediction on data. To achieve that, the input data is employed to construct a model so that a data-driven decision can be made with various static program instructions (Fuyan, 2005). The machine learning algorithm can be broadly categorized into supervised learning, unsupervised, and semi-supervised learning. Supervised learning (input observation mapped with output observation) is learning where the input observation consists of features and the output observation consists of labels (Hastie, Tibshirani, & Friedman, 2009).

Thus, it constructs a model by utilizing a labelled dataset as input (Mohri, Rostamizadeh, & Talwalkar, 2012) and produces a labeled output data. The primary purpose of supervised learning is to drive a functional correlation from the training data with well-generalized testing data. Some of the examples of supervised learning algorithm are employed in classification and regression problems which includes Naïve Bayes, Decision tree, and Logistic regression. Unsupervised learning, on the other hand, is a learning algorithm that is employed when there are difficulties in finding the labeled sample since it does not rely on the previous training for mining the data. The primary purpose of unsupervised learning is to find a correlation that exists between the samples behind the observation. One of the notable examples of unsupervised learning is a clustering system. The semi-supervised learning is a combination of supervised and unsupervised learning that uses a small amount of labelled data and a huge amount of unlabeled data (Tsur, Davidov, & Rappoport, 2010) during the training process. Information recommendation systems and semi-supervised classification are examples of a semi-supervised learning algorithm.

The machine (and also deep) learning algorithm can be applied in many fields of research which include natural language processing (Oyelade and Ezugwu, 2020), medical diagnosis, financial data analysis, bioinformatics, and video surveillance. In the next section, we present our approach of harmonizing machine learning algorithms and IoT technologies using a novel framework. Furthermore, a detailed discussion on the applicability of the proposed framework in combating the novel coronavirus disease is presented in Section 4.

## 3. Proposed Method

The ubiquitous nature of internet-connected sensory devices which often capable of generating relevant data for analytics purpose have motivated the approach promoted in this study. These devices are able to capture structured and unstrucuted data based on time and location of the physical world in high volume. We argue that intelligently processing of these voluminous data requires learnable algorithms which with minimal human intervention can derive a pattern sufficient in presenting higher level information to support combating COVID-19. Hence, in this section, this study presents a novel framework which intelligently allows for interoperability of IoT concepts and machine learning models.

### 3.1 The Smart city based machine learning framework for combating COVID-19

To address the multiple dimensional challenges outlined in Section 1, as a result of COVID-19, a proposed framework is required within the smart city context to allow decision makers to take a crucial decision on the best way to combat COVID-19 from multiple dimensions. The framework consists of multiple components as shown in Figure 1. Each component has a major impact on enhancing the quality of the analytics to combat COVID-19.

**Figure 1:**
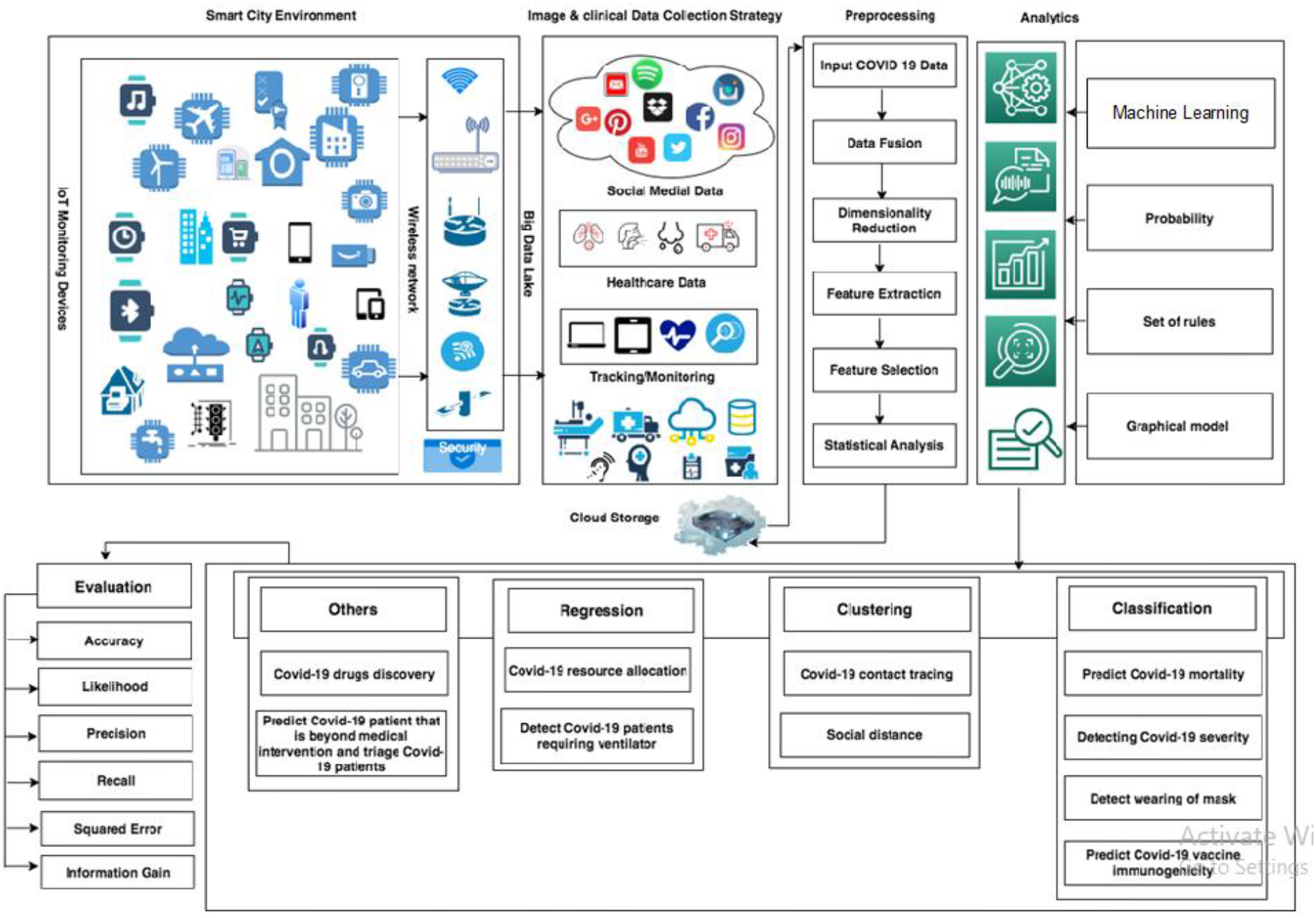
Propose smart city framework integrated with machine learning for fighting COVID-19

### 3.2 Modules of the Smart City Framework

The smart city based framework has four core modules namely: smart city environment, image and clinical collection strategy, image preprocessing, analytics, machine learning models, cloud-based storage, and evaluation strategies. The following sub-sections provide details on some of the modules.

#### 3.2.1 Smart Environment

Smart city technologies have recently demonstrated a major potential in enhancement of citizen’s quality of life. Many smart based technologies have benefited from the adaptation of the internet of things (IoT) by witnessing the development of intelligent applications such as smart home, smart grids, smart transportation, smart industry and smart healthcare. The emphasis on smart technologies during the COVID-19 pandemic could be of great help to tackle major clinical problems and diseases. Moreover, in recent year’s sensors and video cameras surveillance are becoming part of smart city monitoring devices which can lead to an early detection of pandemic. Besides, health agencies may utilize IoT platforms to access data for monitoring the COVID-19 pandemic. For example, ‘Worldometer’ is able to view an instant updates of COVID-19 actual number of cases and deaths for the entire world including daily new cases of COVID-19, distribution of COVID-19 by countries and severity of the disease (Ting et al., 2020). In Figure 1, we demonstrate the smart environment, where various IoT and sensors monitoring devices can possibly be used for healthcare purposes interacting within a limited area to generated numerous clinical signs and symptoms data. These devices are connected via next-generation wireless connectivity which is capable of efficiently transferring the collected data and stored in a big data lake. Big Data plays a critical role in the smart cities because of it is ecosystem of data analytics that can make decision makers take a crucial decision on the best way to further develop strategy to combat COVID-19. With big data, the adequacy of smart city framework implementation, users can be traced at all time with the potential to mitigate any health problems the user may encounter during the movement. Therefore, improve smart city framework efficiency and effectiveness, which in turn improve the life of the citizens living in the smart cities. A literature review was conducted by (Al-Turjman, 2019) the author presents a comprehensive background about 5G standards and it is IoT-specific applications. Furthermore, an overview of recent directions in the utilization of smartphones sensors that can contribute to a scalable operation in smart social spaces is presented.

#### 3.2.2 Image and clinical data collection strategy

The generated image and clinical data from the smart cities in real-time can be subjected to big data processing to better understand healthcare trends, model risk associations and predict outcomes. The result of the big data lake can be used by the government authorities and private/public healthcare providers to improve healthcare services to the citizens, this process will continue until government and healthcare services providers satisfied the citizens living in the smart cities. There are various ways of collecting data at large scale including social media platforms such as (Facebook, Twitter, Google+, Instagram, etc), healthcare services provided through treatment and diagnosis, and tracking monitoring devices such as GPS, vehicle tracking system, smart watches and sensors. All collected data are integrated and stored in a single location within the smart city to be accessed by the authorized entities. The possible technologies used to store such data are Hadoop distributed file system (HDFS) and NoSQL where structured and unstructured data can be stored and processed. In a study by Balduini et al. (2019), the authors proposed a new conceptual framework that put in use a variation of Big Data sources. The framework has a unified approach that make use of spatial and temporal analysis on a heterogeneous stream of data. Based on the results, the study shows generality, feasibility, and effectiveness of the proposed framework through many use cases and examples obtained from real-world requirements collected in many cities.

#### 3.2.3 Pre-processing

In order to provide an accurate and better input for more reliable results to detect and prevent COVID-19 cases, data pre-processing is considered an important stage. The first steps in pre-processing is to extract all the relevant COVID-19 data from the storage. The second step is to preform data fusion where the collected data are integrated to produce more consistent, accurate, and useful information. The third step during pre-processing of COVID-19 data is to preform dimensionality reduction, in which the number of variables are reduced by extracting a set of main variables. Fourth and fifth steps focus on feature extraction and selection. The two methods are very important because it can be used to filter irrelevant or redundant features from the selected datasets. The possible activities in these steps including Wrappers, Filters, and Embedded. The last step is a basic statistical analysis on COVID-19 data in order to interpret the data before intelligent based algorithms are applied.

#### 3.2.4 Analytics

Recent years have received an increasing trend in image processing in healthcare due to convolutional neural network which has become a significant approach for handling large amount of images generated from the smart cities. Allam and Jones (2020) discuss the universal data sharing standards coupled with AI to benefit urban health monitoring and management. The proposed framework has the potential to incorporate various machine learning and deep learning algorithms to develop the analytical model. These algorithms range from traditional shallow approach such as neural network, Decision Tree, Naive Bayes, and K-nearest neighbor. These algorithms can be applied to run on COVID-19 dataset to help in the combatting COVID-19 in hospitals in smart cities across the world. These applications include detect, prevent the spread of COVID-19, forecast next epidemic, diagnose cases, monitor COVID-19 patient, suggest vaccine development, track potential patient, help COVID-19 drug discovery and provide better understand of the COVID-19 virus in smart cities.

##### A. Social media information verification

The COVID-19 pandemic has come with the challenges of fake news associated with it including conspiracy theories. Since the COVID-19 pandemic start spreading across the globe, there has been a lot of fake news regarding the COVID-19 origin, cure, mode of spread, treatment, and many other myths. This is very common on the social media platforms such as Facebook, Twitter, Instagram, YouTube, etc. In the smart city, citizens voice out opinion on social media regarding COVID-19, as such generate a lot of unstructured data. It is reported by (Obeidat, 2020) that no systematic quantitative study has been conducted to ascertain the magnitude of the myths regarding COVID-19 on the social media but certainly the figures of the misinformation about COVID-19 is significant. The fake news regarding COVID-19 can come in the form of manipulated content, misleading content, satire, false context, malicious account, fabricated content, false connection and imposter content. Therefore, machine learning or deep learning algorithms can be applied to detect fake news regarding COVID-19 on the social media and alerts the citizens living in smart cities.

##### B. Prediction of Future pandemic

Although being the worst phases in recent times, this pandemic has come in the times of digital age. Therefore, every aspect of analysis is now being captured including macro-level analysis, logistics, biological etc., in terms of data which will definitely be fruitful for predicting the new and unknown (possibly sources) future pandemic. For example, with the help of machine learning approach (random forest), (Eng, Tong, & Tan, 2014) were able to predict the possible zoonotic strains of influenza i.e. some virus which could only affect animals can also be dangerous to humans. It only implies that machine learning can help in predicting future pandemic which may come from any species. The only limitation being the data could be from different domain i.e. the source of COVID-19 is possibly from ‘bats’ and in the past, sources of pandemic are different (say, different gnome structure etc.). And traditional machine learning requires data distribution to be from the same domains in training and testing. However, Transfer Learning (TL), a part of machine learning, can effectively handle such situations where training and testing data could be from different data distribution. That is, the knowledge learned from the past pandemics could be used in other situations with new domain even with fewer amounts of data. Such scenario is shown in Figure 2, where the pre-trained model from current COVID-19 pandemic (with large data and labels) could be used to predict future pandemics, prepare the smart city for that situation and help overcome quickly in case of the spread (with very less amount of data and labels).

**Figure 2:**
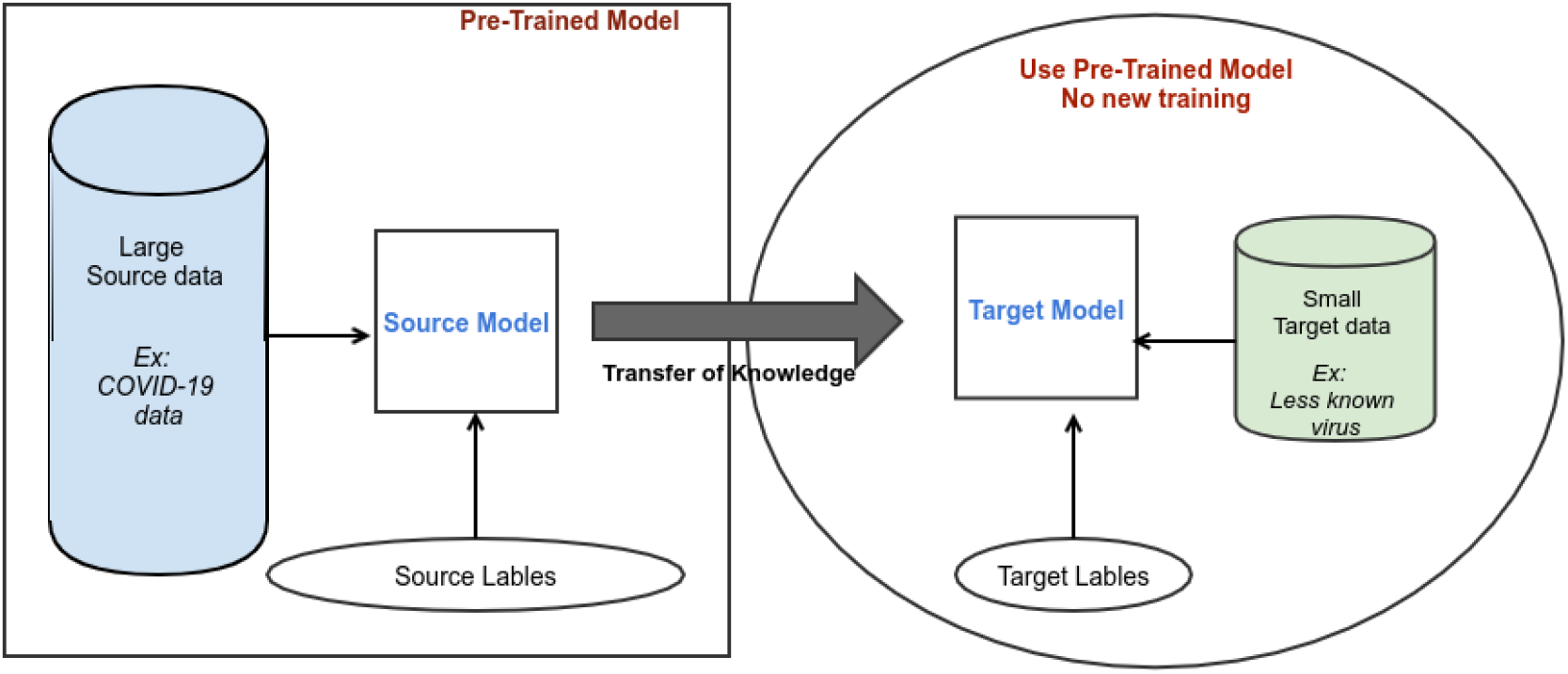
Typical transfer learning model for future pandemic predictions

It is interesting to note that whereas the modules of the smart city based framework for combating COVID-19 presents a very promising applicability, this study further provides perspective on how this is achievable. The following section is focused in detailing on this concept.

## 4. Perspective on the Applicability of the proposed framework

Smart city being an inter-connected urban society and collecting data every moment from several embedded devices, smart cities can effectively work with machine learning approaches during this COVID-19 pandemic. It basically implies that machine learning techniques are hugely dependent on data for better learning and predictive models. Thus, the machine learning techniques have the ability to bring out some intrinsic and useful insights to help smart cities decision makers to take preventive measures during the COVID-19 pandemic. In view of the fact that different machine learning techniques interoperates with other fields of Artificial intelligence (AI), which is the model with the ability to self-learn, this provide a rich self-learn platform. It is important to discuss the role of the AI along with the machine learning to combat COVID-19 because first of all the data availability is too limited and we have to deal with real-time streaming of the data. Thus, significance of self-learning systems becomes much more desirable in smart cities. Figure 3 demonstrates the overall flow of how AI & machine learning approaches can help in fighting COVID-19 pandemic in smart cities.

**Figure 3:**
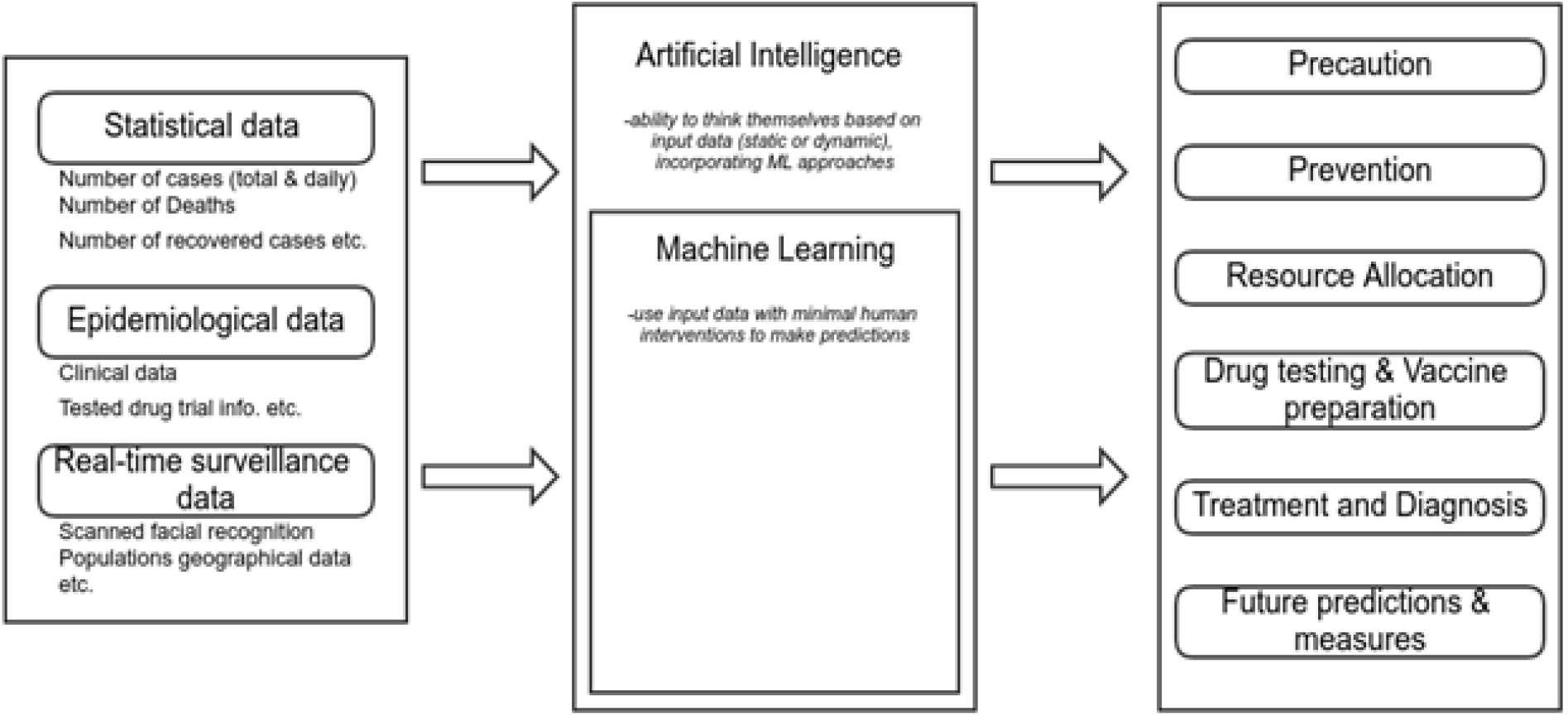
Application and flow of machine learning and other sub-domains of AI in combatting COVID-19

As shown in Figure 3, there are several types of data generated from the information and communication technology equipment embedded in the smart cities. These data are as follows:

i. First kind of data is the statistical data that normally contains the daily statistics of number of identified cases, no of positive cases, number of deaths, number of recovered cases, etc. This would help in the prediction of future cases to prepare for emergencies.
ii. Second type of data is the epidemiological data, which majorly contains all of the clinical tests data concerning test results of different medication, various drug trails, patient’s medical history, patient’s response to several medications, etc.
iii. Third type of data is the real-time surveillance data generated from sensors and cameras in the smart cities. One of the initial detection of COVID-19 is based on symptoms is the fever. People body temperature as per facial recognition and other personal information are the type of data which are also useful to prevent spreading of COVID-19.

The data is processed and analyzed through the machine learning approaches for extracting insights in various applications. The applications of the machine learning in different aspect for combatting COVID-19 are discussed as follows:

### 4.1 Prevention and precaution

Based on the statistical data, machine learning model can be used to predict the nature of the identified cases to better take the preventive measures. During the pandemic situation, there is a chaos and quick testing of individuals on the large scale which is very challenging. Rather than going door to door to each patient, even a less accurate but faster approach is much more acceptable. Machine learning may help in quickly diagnosing the patients in the smart cities as follows:

i. Facial recognitions with the help of sensors and cameras to scan the patients for body temperature and personal information so that if the particular patient is positive then his/her and his/her nearby individuals can be tested and alarmed.
ii. Helping the patients with the AI powered chatbots for self-awareness and answering the queries which might be impossible for the medical professional to address during this COVID-19 because of very high number of patients.
iii. Use the data from the smartphones and wearable smart watches to monitor the heart rate and daily activity of the citizens.

Although, predictions based on the statistical data may not be 100 % accurate, but they can still enable the decision makers in the smart cities to take some preventive and proactive measures.

### 4.2 Prediction models

Medical science (especially dermatology) was one of the real-world fields where AI and machine learning approaches were successfully implemented. Computer vision and machine learning prediction models are enforced for identifying most of the diseases common in the patients by just learning from the images. In case of COVID-19, based on some set of crucial features (set of symptoms), machine learning approaches can help in predicting the following cases:

i. Prediction of a person infected with COVID-19.
ii. Prediction of a positively diagnosed COVID-19 patient to be hospitalized.
iii. Scope of certain treatments to be effective while on treatment. This also includes predicting the chances of a COVID-19 patient being successfully cured or not survives at all.

(Pourhomayoun & Shakibi, 2020) used the machine learning techniques to predict the mortality rate of the patients infected with COVID-19 disease. They used the machine learning algorithms such as: random forest, logistic regression, decision tree, support vector machines, artificial neural networks etc. to predict up to 93% total accuracy in the prediction of the mortality rate. Moreover, the study also used the machine learning models to extract the essential and exclusive symptoms and features to detect the virus.

#### 4.2.1 Prediction of COVID-19 pandemic

Different studies have been conducted to predict the likely occurrence of the COVID-19 pandemic (Arkes, 2001).For instance, (Ndiaye, Tendeng, & Seck, 2020) conducted a prediction study on COVID-19 pandemic globally between January and April 2020. The study employed prophet (Taylor & Letham, 2017), a tool for predicting time series data, which depends on the additive model that fits real non-linear trends with daily, weekly, and annual seasonality together with holiday effects. Four countries involving Italy, China, Senegal, and Iran were selected as case studies for the research. However, the predictive performance of the study shows that the COVID-19 pandemic in some countries like China under the optimistic estimation will end in few weeks whereas the strike of the anti-pandemic in other countries of the world like Italy, Senegal and Iran will end within the end of April. Similarly, (Wang & Wong, 2020) proposed a COVID-Net, using a convolutional neural network design for the identification of COVID-19 cases using chest X-ray (CXR) images. The study utilized the CXR dataset that comprised of 13,800 chest radiography images obtained from 13,725 patients from three public datasets. The experimental analysis shows that the proposed COVID-Net attained a predictive accuracy of 92.6% on the test data, which indicated the importance of combining human and machine collaborative design strategy for building modified deep neural network architectures in a faster mood fitted around the data, task and working requirements.

In another study, (Yang et al., 2020) proposed a modified Susceptible Exposed Infections Removed (SEIR) and AI prediction of COVID-19 pandemics. The study employed the most updated COVID-19 epidemiological data together with population migration data obtained prior and after January 23, 2020, into the SEIR model. Besides, a machine learning approach was employed to train on the 2003 SARS data for the pandemic prediction. The predictive result of the study shows that the pandemic of China is expected to be at peak by late February, which shows a gradual decline by the end of April. However, the cases of the pandemic would have risen higher than expected in Mainland China should the implementation of the proposed model had delayed for 5 days. Thus, the proposed model was effective in predicting the peaks and sizes of the COVID-19 pandemic and the implementation of the control precaution performed on 23rd January was important in alleviating the chances of COVID-19 pandemic size.

Progressively, (Gozes et al., 2020) in their study on the outbreak of COVID-19, developed an artificial intelligence-based automated computer Tomography (CT) image analysis tool using a deep learning approach for the detection, tracking, and quantification of COVID-19, which can identify patient infected with COVID-19 and those that were not infected. The study utilized various global datasets that included Chinese disease-infected areas. Various retrospective deep learning experiment was performed to analyze the system performance in the identification of speculated thoracic Computer Tomography features of the COVID-19 for the evaluation of disease evolution in every patient. One hundred and fifty-seven (157) patients were globally selected from the US and China for the testing sets. However, the classification performance of the model attained 0.996% AUC, 92.2% specificity, and 98.2% sensitivity on Chinese control and infected patient datasets. This shows that the proposed model can attain high predictive performance in the identification, tracking, and quantification of the COVID-19 pandemic. Similarly, (Narin, Kaya, & Pamuk, 2020) on their proposal for automatic prediction of COVID-19, employed a deep convolutional neural network built on Chest X-ray image and pre-trained transfer model that includes InceptionV3, Inception-ResNetV2 and ResNet50 models to attain higher predictive performance with negligible X-ray dataset. However, the experimental results attained an optimum result of 98% accuracy on ResNet50 pre-trained model out of the three selected models, which shows that the research can assist doctors on decision making in clinical practice based on the obtained result as it can employ transfer learning to detect the early stage of the COVID-19 on the infected patients.

#### 4.2.2 Forecasting of Mortality

Since the outbreak of the COVID-19 at Wuhan city China in December 2019, the number of confirmed dead has risen to over 115,000 deaths, which leads to the doubling of the number of deaths on weekly basis (Brown, Jha, & Consortium, 2020). There has been less bias in mortality compared with the reporting of the cases, which might have been influenced by the test policies. Thus, the daily report of deaths on COVID-19 has been on variance with the actual deaths over time. Accurate death rate estimation is important as it serves as a key factor in concluding if a highly infectious disease is seen as a public concern. Consequently, there is a need for a reliable number of mortality estimates on COVID-19, the peak of deaths date, and the period of high mortality to assist in response towards the present and unforeseen pandemics. (Brown et al., 2020) developed a statistical model; also known as Global-19 Assessment of mortality trends in 12 countries between April 12, 2020, and October 1, 2020. The selected countries include the USA, China (Hubei), Italy, Spain, France, UK, Belgium, Iran, Netherlands, Germany, Canada, and Switzerland. Besides, six US states that include New York, Michigan, California, New Jersey, Washington, and Louisiana were incorporated in the study. The mortality data were collected from the WHO country’s daily reports and some obtained online data collection. The results of the prediction in the selected countries showed the estimation of the peak date of the mortality and indicated that the pandemic will be completely wiped off by July 1, 2020. Thus, the model validation provided a reasonable performance with real counts of deaths in various countries. Similarly, Wang et al. (2020) employed the Patient Information Based Algorithm (PIBA) to determine the mortality rate on the COVID-19 pandemic. The algorithm is employed to determine the death rate of the newly infected disease in real-time and forecast future deaths. The data was collected from the 3 public sites of COVID-19 patients in China. The data consists of the newly infected patients with the COVID-19, the patients that died of the infection, the patients with the critical condition and were admitted to the intensive care unit (ICU), daily new cases of the people infected by COVID-19, people with close contacts to the source of infection, and new deaths. The results of the findings show that the average time frame from the beginning of the infection to the time of death is 13 days. This death rate prediction is based on the data collected from Wuhan, the first confirmed city that recorded various death cases related to COVID-19 pandemic.

### 4.3 Resource Allocation

Resources required to manage COVID-19 pandemic become scarce as a result of very high number of people demanding such resources. These resources range from ventilators mask, testing kids, personal protection equipment, sanitizers, etc. The problem of resource allocation is a NP-Hard problem and it is impossible to solve in a polynomial time. Considering the emergency during COVID-19 pandemic situation, machine learning can be very beneficial in the resource allocation prediction on the basis of linear and logistic regression. Even on the small set of training dataset (as is the case in COVID-19), machine learning model may provide feasibility and resource allocation accuracy as close as possible to the optimal resource allocation.

### 4.4 Vaccine discovery

The process of discovering new vaccine based on the available clinical data may take a longer time. But with the help of machine learning approaches, the overall process can be reduced significantly without sacrificing the quality of the vaccine. For example, researcher used the Bayesian machine learning model in a study to discover vaccine for Ebola (Ekins et al., 2015). Also, (Zhang et al., 2017) used the random forest for improving the accuracy of the scores while working on the H7N9. However, even if the vaccine will take time to be found, machine learning can help in finding the existing drugs which may be effective in dealing with COVID-19. Machine learning can learn from drug and protein structure and predict the integrations to generate the studies. Currently, there are different efforts from the scientific community applying machine learning to search for COVID-19 vaccine. (Gonzalez-Dias et al., 2020) reported the stages of applying machine learning to predict signature of vaccine immunogenicity and reactogenicity. The stages involve: data preparation, vaccines and relevant gene selection, selecting the suitable machine learning algorithm for modeling and lastly performance evaluation of the predictive model.

### 4.5 Drug discovery for COVID-19

Since the COVID-19 pandemic, it has become necessary to find the right drugs that can be employed to cure the disease. Various approaches have been attempted in finding the right drugs by either repurposing the existing one (Therapeutic) or discovering the new one. The application of machine learning and the development of the new models have made researchers focus on the application of machine and deep learning models for the discovery of drugs that can bring a cure for COVID-19. A review of some of the studies that applied the machine or deep learning approach for drug discovery and vaccine for COVID-19 are summarized in Table 1.

**Table 1:** Summary of the application of machine learning in drugs discovery

| Reference | Study approach | Advantage | Challenge |
| --- | --- | --- | --- |
| (Patankar, 2020) | Deep learning-based computational drug discovery | The generated molecules could serve as a good drug potential for SARs CoV and COVID-19. The result also shows that a deep learning model is important in testing the current compound and obtaining a new molecule for finding COVID-19 drugs. | By utilizing a limited data for model training, there could be an atom of error resulting from inaccuracies of parameters. |
| (Chenthamarakshan et al., 2020) | Target-specific and selective drug design for COVID-19 using Deep generative model | The framework can take care of candidate generations for multiple targets by employing trained mode. Besides, it can also permit explicit accounting of off-target selectivity | Large scale data size is required for effective study |

## 5. Results and Discussion

In this section, we present the findings of the study and reinforced the importance of proposed framework by using use cases where machine learning techniques are applied to help in combating COVID-19 in smart cities.

The key outcome of this study is the proposed framework and the wide range of its applicability to the advancement of global efforts in curbing the devastating effect of COVID-19. The findings from this study showed that handcrafted and manually driven mechanisms for managing COVID-19 remains ineffective as the outbreak in all its instances is overwhelming, defying such mechanisms. The pervasive nature of data-driven smart devices and applications continue to provide intelligent and scalable solution for achieving a secure city in an event of endemic and global pandemic as COVID-19. This finding is further supported by Shorfuzzaman et al, (2021) which argued that mass video surveillance provides a great potential for managing social distancing as a panacea to slow down the propagation of the disease. Their study reecho the aim of this study which demonstrates that data-driven driven machine learning frameworks dependent or sensory devices will largely allow for artificial deployment of smart cities in curbing COVID-19. This study also discovered that in addition to the benefit of managing social distancing through surveillance, such video files can also provide contact tracing applications with input in chronicling events and persons needing tracing. In another related study confirming the methods used in this study and its findings and the profitability to cities overwhelmed by COVID-19, Allam et al (2021) noted that 6G technology, including Digital Twins and Immersive Realities (XR) will support the socio-economic of its population.

Countries across the globe have had their share of first, second and even third waves of the pandemic and are beginning to look towards a post-pandemic era with digitalization of city-system to manage future outbreak. Again, this concretizes the critique presented in this study which highlights the need for ‘smarting up’ the systems that drives cities. The study of Graziano et al, (2021) supports the potency of the proposed framework in this study. The authors noted that governments are now considering a more inclusive tech-led urban development – smart cities. We argue that in developing such tech-urban settlement, the outcome of this study presents authorities with machine learning driven framework for curtailing and managing subsequent and imminent wave of the disease except a global vaccination is achieved. For instance, in leveraging the real-time data collection through sensory devices in smart cities, studies using machine and deep learning algorithms have built temporal learning algorithms. Sun et al, (2021) supported this claim by applying deep learning, a sub-model of machine learning model, in projecting the level of COVID-19 disease outbreak using temporal data which are richly generated in a smart city driven by our proposed framework. On the imaging and preprocessing case proposed by the framework in this study, Lassau et al, (2021) showed that by intelligently integrating the performance of deep learning models with related variables (e.g clinical and biological), the severity of the disease can be predicted ahead of time in patients. Again, adoption of the proposed framework in this study presents city premiers with a potent tool for managing future events. Also, the works of Shortenet al (2021) and Pan et al (2021) demonstrates the importance of integrating machine learning algorithms in city-wide management of COVID-19 as achieved in this study through the proposed framework.

The use case can help readers really understand exactly the way machine learning can provide aid in the era of fighting COVID-19 pandemic. As such, other nations can share the expertise in fighting the COVID-19 by applying the machine learning approaches. The use cases are discussed as follows:

### Case Study: New York City

In New York city, there are heavy cases of COVID-19 patients and those exhibiting the symptoms. The medical staffs in New York hospitals are Overwhelmed in view of the fact that the number of COVID-19 patients is extremely high. As a result of that, the medical staffs are facing difficulties in taken decision on the COVID-19 patient that require emergency treatment and the patient that might have case beyond medical intervention. To speed up the decision of the medical staffs in New York hospitals, machine learning system is developed through training of the system to provide clinical decisions support to triage patients. It is the system that is now in used in the hospitals in aiding clinical decisions (Spectrum, 2020).

### Case Study: China

When it comes to the issue of data, China is well known about it is massive amount of data generated from it is citizens. China installed a network of over 200 million surveillance cameras spread across the country. In addition to video surveillance camera, biometric scanners are installed in doorways of residential complexes. As a form of registration, any resident or person that is leaving the residential building has to scan his face through the biometric scanner at the doorway of the building. After that, the embedded intelligent systems process the data and track the person location through the video surveillance. All the information are stored in a central database in which the machine learning algorithms runs the data to determine the possible social interaction of the person that leaves the residential building (Dingli, 2020).

### Case Study: Canada

Migration of humans across the globe contributed significantly to the spread of the COVID-19 pandemic throughout the world. BlueDot based in Canada applied machine learning and natural language processing for the tracking, recognition and the reporting of the spread of COVID-19 faster than the WHO and center for disease control and prevention in United States of America. It is projected that this machine learning and natural language processing based technology can be leveraged in the future for the prediction of zoonotic infection risk to humans using climate and human activities as variables. The prediction of individual risk profile using the data extracted from social media such as family history and lifestyle as well as clinical, personal and travel data can provide precise and accurate prediction results. However, such technology can trigger privacy concerned (Obeidat, 2020).Similarly, *Virtual healthcare assistant*: the virtual healthcare assistant is a multi-lingual healthcare agent that is developed based on natural language processing. It is a question – answering system that respond to questions related to COVID-19 by delivering information that is trustworthy on COVID-19 guidelines, protection measures, symptoms monitoring and checking as well as providing advised to individuals on the need for screening in the hospital or self-isolation. The healthcare agent is developed by Canada-based stallion (Obeidat, 2020).

### Case Study: United States of America

In United States, many medical centers are modifying their existing intelligent system that were purposely meant to predict course of patients’ illness. These intelligent systems are now being modified to predict specific type of COVID-19 outcomes like the intubation. The intelligent systems are trained to learned pattern about the illness by feeding the system with thousands of patient records as training data. However, no sufficient data to build entirely new intelligent systems for the prediction of COVID-19. Therefore, researchers are assessing the existing tools with the aim of customizing it to help in the fight against COVID-19 pandemic (Spectrum, 2020).

## 6 Conclusions

In this paper, we propose a solution framework based on machine learning for integration in smart cities to fight COVID-19. The propose machine learning solution framework integrated different task to fight COVID-19 in the smart cities from different dimension such as predicting COVID-19 vaccine immunogenicity, detecting COVID-19 severity, predicting COVID-19 mortality, COVID-19 resource allocation, COVID-19 drugs discovery, COVID-19 contact tracing, social distance, detecting wearing of mask, detecting COVID-19 patients requiring ventilator, predicting COVID-19 patient that is beyond medical intervention and triage COVID-19 patients. The paper presented a comprehensive guide for implementing the machine learning framework in smart cities. The solution framework has the potential to automate the measures of fighting the COVID-19 pandemic in smart cities from multiple dimensions to ease the fatigue of the healthcare workers due to very high number of COVID-19 patients requiring medical attention simultaneously and provide widespread access to quality healthcare system. In addition, this study foresees the proposed smart city machine learning based framework for combatting COVID-19 will serve as essential guide to the research community in developing a more compartmentalized forecasting and analyzing tools with prospect of mitigating the spread of the ongoing COVID-19 pandemic and the reoccurrences of any similar future pandemic disease. The limitation of the study is inherent in the fact that the pieces of the framework is still in its design form and not yet piece-together in implementation form. However, implemented components of the framework, as seen in several studies, already confirms its applicability. For future work, it will be interesting to see the real world application of the proposed framework to further investigates the model practicality and efficiency in smart city environments. This deployment of the proposed framework will generate policies to allow for their effective integration into the existing social and health systems of the city.

## Data Availability

No data is associated with this manuscript

